# Evaluating Changes in Pain Medication Administration During IUD Insertion, 2018-2025

**DOI:** 10.1101/2025.07.17.25331736

**Authors:** Nina B. Masters, Karen M. Gilbert, Brianna M. Goodwin Cartwright, Patricia J. Rodriguez, Duy Do, Nicholas L. Stucky

## Abstract

**Background:** Intrauterine devices (IUDs) are highly effective, long-acting reversible contraceptives that can prevent pregnancy for up to 10 years. IUDs can also provide important non-contraceptive benefits, including reducing heavy periods and cramping, and improving pelvic pain associated with endometriosis. Despite their effectiveness, IUD insertion is painful, and pain management during IUD insertions is uncommon, with most patients just recommended to take NSAIDs such as ibuprofen at home prior to the procedure. The CDC updated their Selected Practice Recommendations for Contraceptive Use in August 2024, including guidance on pain management during IUD insertions.

**Objectives:** This study aims to describe real-world trends in the use of pain medication during IUD insertion over time, specifically evaluating lidocaine and opioid use. We also evaluate the impact of CDC’s 2024 recommendations on pain medication administration during IUD insertion.

**Study Design:** This study used a subset of Truveta Data, a daily updated electronic health record (EHR) database from a collective of US healthcare systems. IUD insertions were included in the study population if the patient was of female biological sex and aged 15-49 years at the time of the procedure and the procedure occurred between January 1, 2018 and June 30, 2025. Lidocaine and opioid medications prescribed or administered on the day of the IUD insertion were captured, and logistic interrupted time series analyses models were used to evaluate predictors associated with receipt of lidocaine or opioids and quantify the impact of the 2024 recommendations.

**Results:** There were 286,144 IUD insertions meeting study criteria between January 1, 2018 and June 30, 2025. IUD insertions were most common in individuals aged 25-39 (40.0%), followed by those aged 35-49 (36.7%) and those aged 15-24 years (23.2%). In 2018, 1.7% of patients received any pain medication with their IUD insertion, 1.2% received lidocaine and 0.8% received an opioid. In 2025, 5.0% of patients received any pain medication; 4.2% received lidocaine and 2.0% received an opioid. This represents a 195% increase in pain medication overall, a 253% increase in lidocaine, and a 142% increase in opioids. After adjusting for other factors in time series models, the CDC guidelines were associated with an 18% immediate increase in the odds of lidocaine receipt (aOR 1.18, 95% CI: 1.05, 1.34) and a 22% immediate increase in the odds of opioid receipt during IUD insertion (aOR 1.22, 95% CI: 1.04, 1.44). However, the monthly trend of pain medication administration and prescriptions for both lidocaine and opioids did not increase significantly after the guidelines.

**Conclusions:** While rates of pain medication administration and prescriptions during IUD procedures have increased nearly threefold since 2018, in 2025, 95% of women receiving an IUD are still not receiving pain medication as part of the procedure.

## Introduction

Intrauterine devices (IUDs) are highly effective, long-acting reversible contraceptives that can prevent pregnancy for between 3 to 10 years based on the IUD type.^1^ IUDs have a very low failure rate,^1^ and their long-term effectiveness makes them a very convenient birth control option. A survey from 2015-2017 showed that 14% of women who use contraceptives aged 15-44 were using an IUD.^2^ For those unable to tolerate oral contraceptive medications, such individuals with migraines or hypertension, IUDs can be an important – and possibly the only – option for contraception.^3^ IUDs can also provide important non-contraceptive benefits, including reducing heavy periods and the severity of cramping, and improving pelvic pain associated with endometriosis.^3^

Despite their effectiveness, IUD insertion is known to be painful, and fear of pain may limit IUDs’ popularity.^4^ A 2015 survey found that 78% of women reported the insertion procedure as moderate to severe, and 46% experienced vasovagal symptoms.^5^ Further, a 2011 study among nulliparous women found that 72% and 17% of patients reported the IUD insertion to be moderately and severely painful, respectively.^6^ Notably, IUD insertion pain is not experienced equally among all groups - factors such as nulliparity (specifically no history of a vaginal delivery)^7,8^, dysmenorrhea^9^, and patient anxiety^10^ have been associated with higher levels of reported pain. Despite known pain during insertion, pain management during IUD insertions is uncommon, and a recent study from the Veteran’s Affairs Health Care System found only 11.4% of IUD insertions were accompanied by pain management prescriptions, of which 8.3% were nonsteroidal anti-inflammatory drugs (NSAIDs), most commonly ibuprofen.^11^ However, multiple randomized trials and systematic reviews have shown that generally, NSAIDs do not significantly reduce IUD insertion-related pain, with the exception of Naproxen.^12,13^ Lidocaine, a local anesthetic, seems more promising in reducing pain during IUD insertion^14^, although different formulations and medication strength impact its effectiveness.^13,15^

In alignment with emerging evidence and increased awareness of patient experiences, including growing public discourse around the pain during IUD insertion that gained visibility on social media platforms like TikTok in 2022,^16,17^ the Centers for Disease Control and Prevention (CDC) updated the US Selected Practice Recommendations for Contraceptive Use in August 2024, including recommendations for pain management during IUD insertions. This document emphasized the importance of discussing and prioritizing pain management with patients, included lidocaine as a preferred option, and recommended against the use of misoprostol.^18,19^ Though not mentioned in the CDC guidelines, opioids, notably tramadol, have been shown to reduce pain with IUD insertion,^13,20^ but are not commonly used during IUD insertions.^11^ This study aims to describe real-world trends in the use of pain medication during IUD insertion over time, specifically evaluating lidocaine and opioid use. We also evaluate the impact of CDC’s 2024 guidelines on pain medication administration during IUD insertion.

## Materials and Methods

### Data

This study used a subset of Truveta Data, a daily updated electronic health record (EHR) database from a collective of US healthcare systems. Data elements used for this analysis include structured demographic information, encounters, conditions/diagnoses, procedures, and medication requests and administration. Data undergo syntactic and semantic normalization. Standard filters on data recency were applied regarding encounter, medication, and procedure data. Truveta Data were de-identified by expert determination under the HIPAA Privacy Rule. This study used only de-identified patient records and therefore did not require Institutional Review Board approval. Data were accessed on July 10, 2025.

### Population

IUD insertions were included in the study population if the patient was of female biological sex and aged 15-49 years at the time of an IUD insertion. IUD insertions between January 1, 2018 and June 30, 2025 were included in the study. IUD insertion was captured with the CPT code “58300” (‘Insertion of intrauterine device (IUD)’). Lidocaine and opioid pain medications prescribed or administered on the day of the IUD insertion procedure were assessed using HCPCS and RxNorm codes (Supplementary Table S1). If a patient received both, both pain medications were included.

Exclusion criteria were applied to remove instances where pain medication may have been prescribed or administered for a different reason than the IUD insertion. First, women who had another gynecological procedure (such as a cervical or endometrial biopsy, colposcopy, etc. based on a procedure list from a systematic review by Ireland and Allen on pain management for office-based gynecologic procedures^21^) on the same day as the IUD insertion were excluded (Supplementary Table S2). Patients who had an IUD insertion in the preceding 3 months were also excluded due to the potential for an expulsion or perforation that may have been responsible for a short interval repeat IUD insertion. Finally, patients with a live birth or who received lidocaine or opioid pain medication in the 30 days prior to the IUD insertion were excluded.

IUD insertions meeting the above criteria (n=303,960) were subset to those with available practitioner details (n = 288,002, 94.7%). Data were then filtered for known practitioner sex or clinician background (removing 0.41% and 0.25% respectively). If multiple providers participated in an encounter, the provider with the highest volume of procedures (IUD insertions) in the population was used.

### Statistical Analysis

Monthly rates of pain medication administration or prescription (lidocaine, opioid, and combined) during IUD insertions were described over time. Interrupted time series analyses leveraging logistic regression were used to explore the impact of the CDC’s August 2024 policy change on pain medication use. Evidence of seasonality and autocorrelation were absent in standard checks, so no seasonal adjustments were made. Statistical analyses included sociodemographic variables that may impact disparities in care including age, race, census region, and ethnicity, in addition to clinical factors that may have impacted pain medication receipt: any prior IUD insertion, prior live birth, prior diagnosis of dysmenorrhea, endometriosis, Polycystic Ovary Syndrome (PCOS), or an anxiety disorder. Practitioner sex and clinician background were also included to evaluate practitioner-level factors. Analyses were conducted in R (version 4.4.1) within a cloud-based notebook environment leveraging Apache Spark, using SparkR.

## Results

### Characteristics of Patients Undergoing IUD Insertions, 2018-2025

There were 286,144 IUD insertions between January 1, 2018 and June 30, 2025, with 243,156 (85.0%) occurring prior to the CDC recommendations in August 2024, and 42,988 (15.0%) occurring after (Table 1). IUD insertions were most common in individuals aged 25-39 (40.0%), followed by those aged 35-49 (36.7%) and those aged 15-24 years (23.2%). Within this study population, 15.6% had evidence of a prior IUD, and 25.3% had evidence of a prior live birth. Comorbidities such as dysmenorrhea, endometriosis, and PCOS were less common, at 10.9%, 2.3%, and 4.3%, respectively. Overall, 2.8% of IUD insertions had pain medications administered or prescribed (2.3% had lidocaine, while 1.3% had opioids).

**Table 1.** Characteristics of Patients Receiving IUD Insertions from January 2018 – June 2025, Stratified by CDC Guideline Update in August 2024.

|  | Overall | Before CDC Guideline | Post CDC Guideline |
| --- | --- | --- | --- |
|  | N = 286,144 (100.0%) | N = 243,156 (85.0%) | N = 42,988 (15.0%) |
| **Age Group** |  |  |  |
| 15-24 | 66,475 (23.2%) | 56,765 (23.3%) | 9,710 (22.6%) |
| 25-34 | 114,533 (40.0%) | 98,406 (40.5%) | 16,127 (37.5%) |
| 35-49 | 105,136 (36.7%) | 87,985 (36.2%) | 17,151 (39.9%) |
| **Race** |  |  |  |
| Asian | 12,022 (4.2%) | 10,093 (4.2%) | 1,929 (4.5%) |
| Black or African American | 23,925 (8.4%) | 20,477 (8.4%) | 3,448 (8.0%) |
| Other | 15,768 (5.5%) | 13,137 (5.4%) | 2,631 (6.1%) |
| White | 205,283 (71.7%) | 176,071 (72.4%) | 29,212 (68.0%) |
| Unknown | 29,146 (10.2%) | 23,378 (9.6%) | 5,768 (13.4%) |
| **Ethnicity** |  |  |  |
| Hispanic or Latino | 26,322 (9.2%) | 21,622 (8.9%) | 4,700 (10.9%) |
| Not Hispanic or Latino | 226,610 (79.2%) | 194,181 (79.9%) | 32,429 (75.4%) |
| Other/Unknown | 33,212 (11.6%) | 27,353 (11.2%) | 5,859 (13.6%) |
| **Census Region** |  |  |  |
| Midwest | 101,725 (35.6%) | 88,092 (36.2%) | 13,633 (31.7%) |
| West | 91,671 (32.0%) | 76,803 (31.6%) | 14,868 (34.6%) |
| South | 57,763 (20.2%) | 48,119 (19.8%) | 9,644 (22.4%) |
| Northeast | 19,034 (6.7%) | 15,921 (6.5%) | 3,113 (7.2%) |
| Unknown | 15,951 (5.6%) | 14,221 (5.8%) | 1,730 (4.0%) |
| **Urbanicity** |  |  |  |
| Rural | 38,702 (13.5%) | 33,182 (13.6%) | 5,520 (12.8%) |
| Urban | 234,517 (82.0%) | 198,449 (81.6%) | 36,068 (83.9%) |
| Unknown | 12,925 (4.5%) | 11,525 (4.7%) | 1,400 (3.3%) |
| **Had an Outpatient Visit in Past Year** | 263,499 (92.1%) | 223,743 (92.0%) | 39,756 (92.5%) |
| **Had a Prior IUD** | 44,733 (15.6%) | 36,742 (15.1%) | 7,991 (18.6%) |
| **Had a Prior Live Birth** | 72,434 (25.3%) | 61,477 (25.3%) | 10,957 (25.5%) |
| **History of Dysmenorrhea** | 31,141 (10.9%) | 25,886 (10.6%) | 5,255 (12.2%) |
| **History of Endometriosis** | 6,590 (2.3%) | 5,388 (2.2%) | 1,202 (2.8%) |
| **History of PCOS** | 12,382 (4.3%) | 9,937 (4.1%) | 2,445 (5.7%) |
| **Received Any Pain Med during IUD Insertion** | 7,953 (2.8%) | 5,959 (2.5%) | 1,994 (4.6%) |
| **Received Lidocaine during IUD Insertion** | 6,629 (2.3%) | 4,918 (2.0%) | 1,711 (4.0%) |
| **Received Opioid during IUD Insertion** | 3,738 (1.3%) | 2,876 (1.2%) | 862 (2.0%) |
| **Practitioner Sex** |  |  |  |
| Female | 224,872 (78.6%) | 190,458 (78.3%) | 34,414 (80.1%) |
| Male | 61,272 (21.4%) | 52,698 (21.7%) | 8,574 (19.9%) |
| **Practitioner Clinical Background** |  |  |  |
| General Practitioner | 65,707 (23.0%) | 56,028 (23.0%) | 9,679 (22.5%) |
| Medical Trainee | 2,880 (1.0%) | 1,761 (0.7%) | 1,119 (2.6%) |
| Midwife | 8,170 (2.9%) | 7,035 (2.9%) | 1,135 (2.6%) |
| Nurse Practitioner/Registered Nurse | 33,027 (11.5%) | 27,737 (11.4%) | 5,290 (12.3%) |
| Obstetrician/Gynecologist | 165,752 (57.9%) | 141,644 (58.3%) | 24,108 (56.1%) |
| Physician's Assistant | 6,981 (2.4%) | 6,074 (2.5%) | 907 (2.1%) |
| Pediatrics | 3,627 (1.3%) | 2,877 (1.2%) | 750 (1.7%) |

### Rate of Pain Medication Administration or Prescription During IUD Insertions

Pain medication administration or prescription was lowest in 2018 and increased over time (Figure 1). In 2018, 1.7% of patients were administered or prescribed pain medication with their IUD insertion; 1.2% received lidocaine and 0.8% received an opioid (Table S3). In 2025, 5.0% of patients were administered or prescribed pain medication with their IUD insertion; 4.2% received lidocaine and 2.0% received an opioid. This represents a 195% increase in pain medication overall, a 253% increase in lidocaine administration, and a 142% increase in opioid administration (Table S3). Pain medication stratified by age group and year is shown in Table S4.

**Figure 1.**
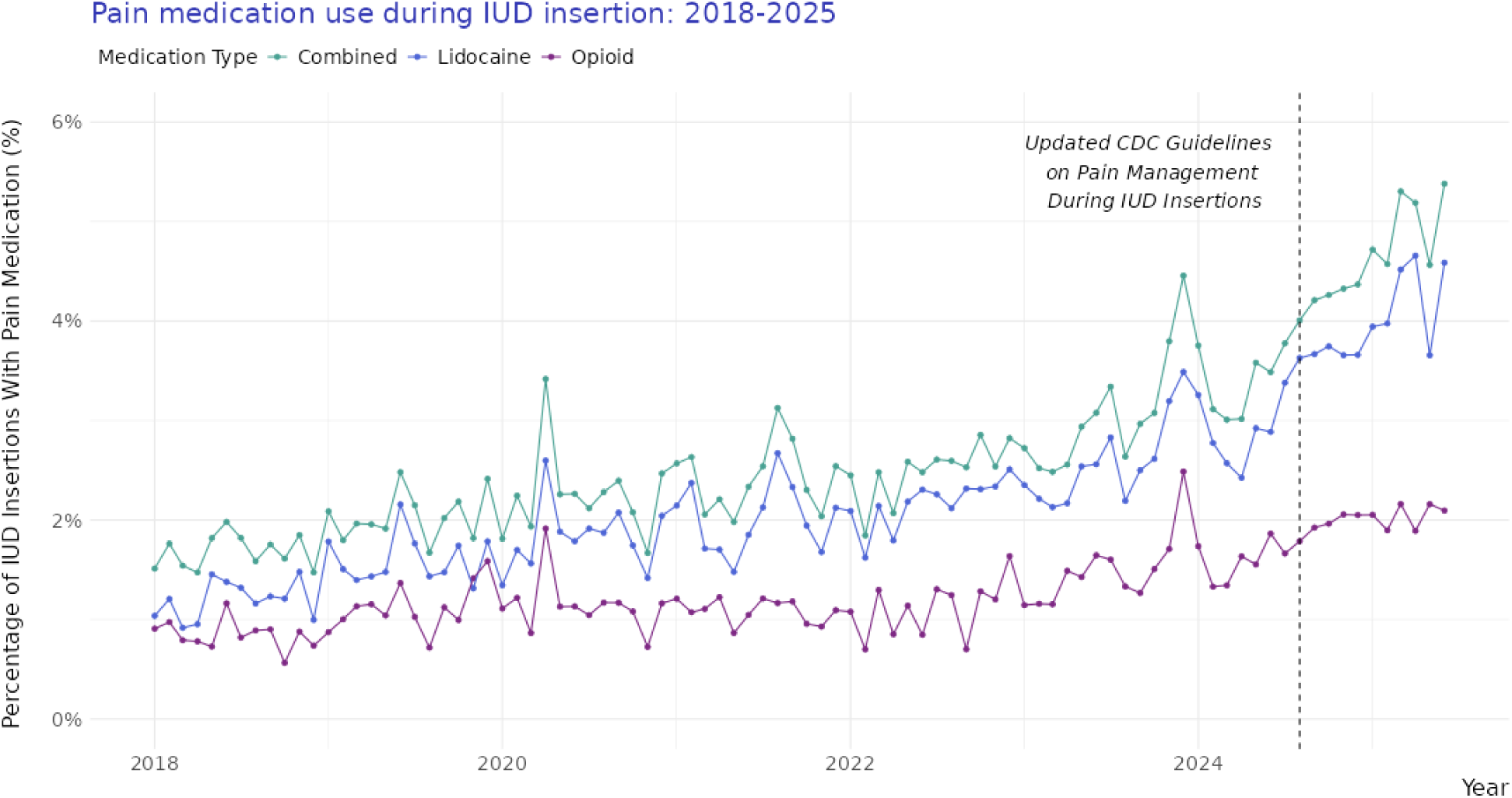
Lidocaine, Opioid, and Overall Pain Medication Administration and Prescription Rates during IUD Insertions, 2018-2025

### Impact of CDC Guidelines on Lidocaine and Opioid Administration or Prescription during IUD Insertions

From January 2018 to August 2024, the odds of lidocaine receipt during IUD insertion increased by 1% each month (aOR 1.01, 95% CI: 1.01, 1.01) (Figure 2, Table 2). After the August 2024 CDC guidelines on pain medication management during IUD insertions, there was an 18% immediate increase in the odds of administering or prescribing lidocaine during IUD insertion (aOR 1.18, 95% CI: 1.05, 1.34). However, there was no additional monthly change in the rate of lidocaine receipt after these guidelines were released (aOR 1.01, 95% CI: 1.00, 1.03). There was a 0.8% monthly increase in the odds of opioid administration or prescription over the study period, with a 22% immediate increase after the CDC guidelines were released (aOR 1.22, 95% CI: 1.04, 1.45) and no trend effect (aOR 1.01, 95% CI: 0.98, 1.03). Counterfactual trajectories based on model outputs are shown in Figure 2.

**Figure 2.**
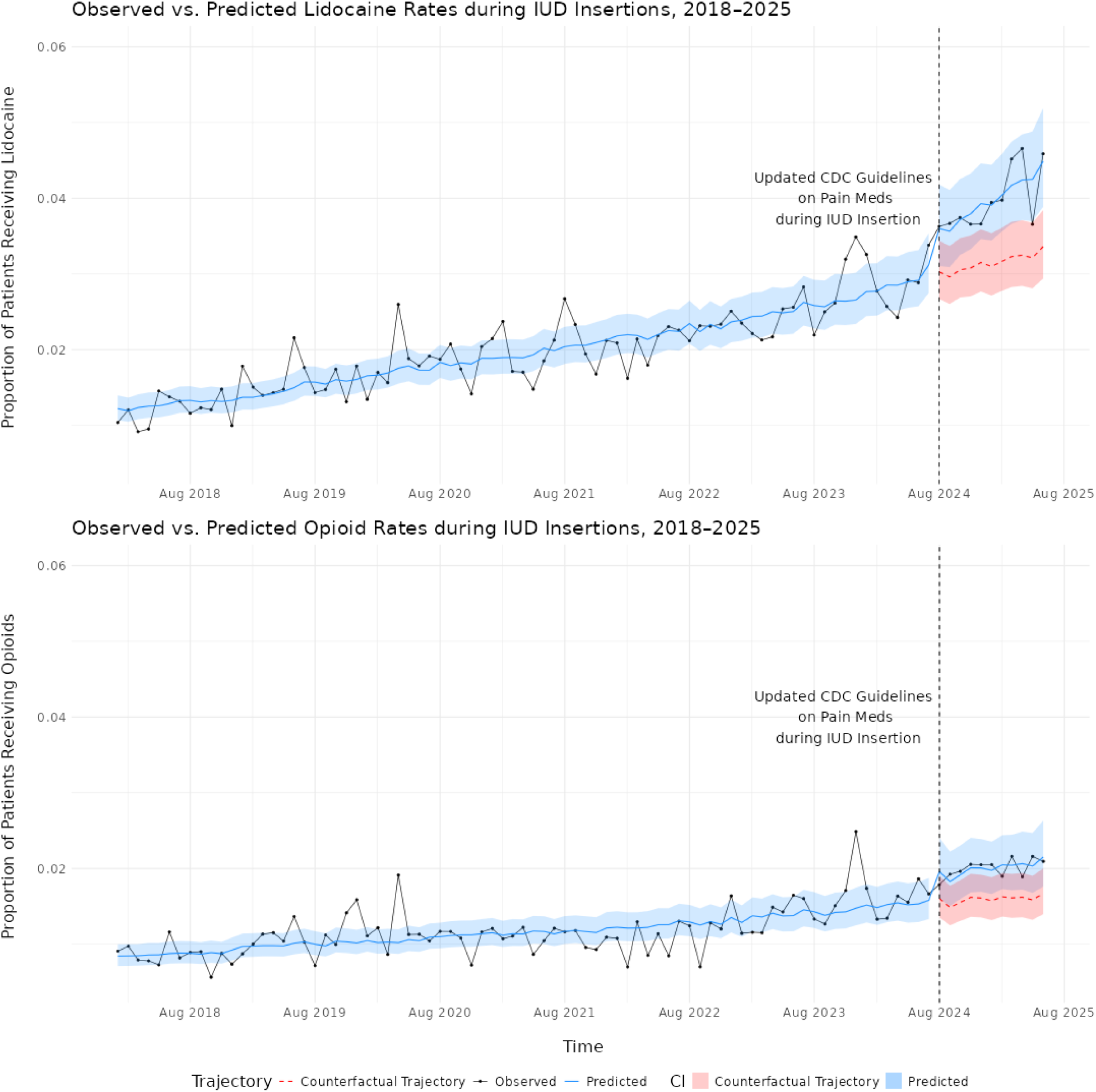
Model of Predicted vs. Counterfactual Trajectory of Lidocaine and Opioid Medication Rates during IUD Insertions, 2018-2025

**Table 2.** Odds of Lidocaine or Opioid Receipt during IUD Insertions: Logistic Regression Interrupted Time Series Models.

| Variable | Lidocaine |  | Opioid |  |
| --- | --- | --- | --- | --- |
|  | Odds Ratio | 95% CI | Odds Ratio | 95% CI |
| **Time Series Variables** |  |  |  |  |
| Month (2018-Aug 2024) | 1.01 | 1.01, 1.01 | 1.01 | 1.01, 1.01 |
| CDC Guideline Update (Aug 2024)* | 1.18 | 1.05, 1.34 | 1.22 | 1.04, 1.44 |
| Month Post CDC Guideline Update ** | 1.01 | 1.00, 1.03 | 1.01 | 0.98, 1.03 |
| **Age Group** |  |  |  |  |
| 15-24 Years | REF | REF | REF | REF |
| 25-34 Years | 0.76 | 0.72, 0.81 | 0.88 | 0.81, 0.95 |
| 35-49 Years | 0.51 | 0.48, 0.55 | 0.52 | 0.47, 0.56 |
| **Race** |  |  |  |  |
| White | REF | REF | REF | REF |
| Asian | 0.90 | 0.79, 1.03 | 1.05 | 0.86, 1.28 |
| Black | 1.69 | 1.56, 1.84 | 2.71 | 2.47, 2.96 |
| Other*** | 1.20 | 1.07, 1.33 | 1.09 | 0.92, 1.29 |
| No Information | 1.18 | 1.07, 1.29 | 1.13 | 0.99, 1.29 |
| **Ethnicity** |  |  |  |  |
| Hispanic or Latino | REF | REF | REF | REF |
| Non Hispanic or Latino | 0.95 | 0.86, 1.04 | 0.81 | 0.72, 0.91 |
| Other/Unknown | 0.94 | 0.84, 1.05 | 0.74 | 0.63, 0.87 |
| **Census Region** |  |  |  |  |
| Midwest | REF | REF | REF | REF |
| West | 2.73 | 2.56, 2.92 | 0.84 | 0.77, 0.92 |
| South | 1.41 | 1.31, 1.52 | 0.72 | 0.66, 0.79 |
| Northeast | 0.96 | 0.85, 1.09 | 0.86 | 0.76, 0.98 |
| Unknown | 0.64 | 0.51, 0.81 | 0.88 | 0.69, 1.12 |
| **Urbanicity** |  |  |  |  |
| Rural | REF | REF | REF | REF |
| Urban | 1.48 | 1.35, 1.62 | 1.08 | 0.96, 1.21 |
| Unknown | 2.42 | 1.96, 3.00 | 1.47 | 1.16, 1.88 |
| **Clinical Factors^** |  |  |  |  |
| Prior IUD | 0.99 | 0.92, 1.06 | 0.82 | 0.74, 0.90 |
| Prior Live Birth | 0.86 | 0.81, 0.91 | 0.57 | 0.52, 0.62 |
| Prior Dysmenorrhea Diagnosis | 1.42 | 1.33, 1.51 | 1.46 | 1.33, 1.59 |
| Prior Endometriosis Diagnosis | 1.98 | 1.76, 2.22 | 2.33 | 2.02, 2.69 |
| Prior PCOS Diagnosis | 1.16 | 1.04, 1.29 | 1.04 | 0.90, 1.21 |
| Prior Anxiety Diagnosis | 1.25 | 1.19, 1.32 | 1.32 | 1.23, 1.41 |
| **Practitioner Sex** |  |  |  |  |
| Female | REF | REF | REF | REF |
| Male | 0.90 | 0.84, 0.95 | 0.93 | 0.86, 1.01 |
| **Clinician Background** |  |  |  |  |
| General Practitioner | REF | REF | REF | REF |
| OBGYN | 1.90 | 1.78, 2.04 | 3.48 | 3.11, 3.90 |
| Pediatrics | 1.14 | 0.92, 1.41 | 0.74 | 0.47, 1.15 |
| NP/RN | 1.23 | 1.12, 1.36 | 0.44 | 0.35, 0.56 |
| Midwife | 0.34 | 0.25, 0.45 | 0.52 | 0.34, 0.79 |
| PA | 2.06 | 1.77, 2.40 | 0.67 | 0.45, 1.02 |
| Medical Trainee | 2.15 | 1.79, 2.60 | 2.20 | 1.62, 2.99 |

\* CDC Guideline Update is a binary variable indicating that IUD insertion occurred on or after August 2024
\*\*Month post CDC Guideline Update refers to months elapsed since August 2024
\*\*\*Other race includes multiple races and single races representing less than 2% of the overall study population
^Reference for each group is the absence of the condition

### Patient-Level Predictors of Receipt of Lidocaine or Opioid during IUD Insertions

Patient-level factors from separate models for lidocaine and opioid receipt outcomes are shown in Figure 3 (Table 2). Receipt of lidocaine and opioids varied by age group, with women aged 35-49 and 25-34 significantly less likely to receive lidocaine (aOR 0.51, [95% CI: 0.48, 0.55], aOR 0.76, [95% CI: 0.72, 0.81]) and opioids (aOR 0.52, [95% CI: 0.47, 0.56], aOR 0.88, [95% CI: 0.81, 0.95]) than women aged 15-24. Patients residing in urban areas had greater odds of receiving lidocaine (aOR 1.48, [95% CI: 1.35, 1.62]) than those in rural areas. Black patients had elevated odds of receiving both lidocaine (aOR 1.69, 95% CI: 1.56, 1.84) and opioids (aOR 2.71, [95% CI: 2.47, 2.96]) compared to White patients. Patients with comorbid conditions causing pelvic pain generally had increased odds of receipt of pain medication. Specifically, patients with endometriosis had increased odds of receiving both lidocaine (aOR 1.98, [95% CI: 1.76, 2.22]) and opioids (aOR 2.33, [95% CI: 2.02, 2.69]). Women with a prior live birth had decreased odds of receiving lidocaine (aOR 0.86, [95% CI: 0.81, 0.91]) and opioids (aOR 0.57, [95% CI: 0.52, 0.62]) compared to nulliparous women.

**Figure 3.**
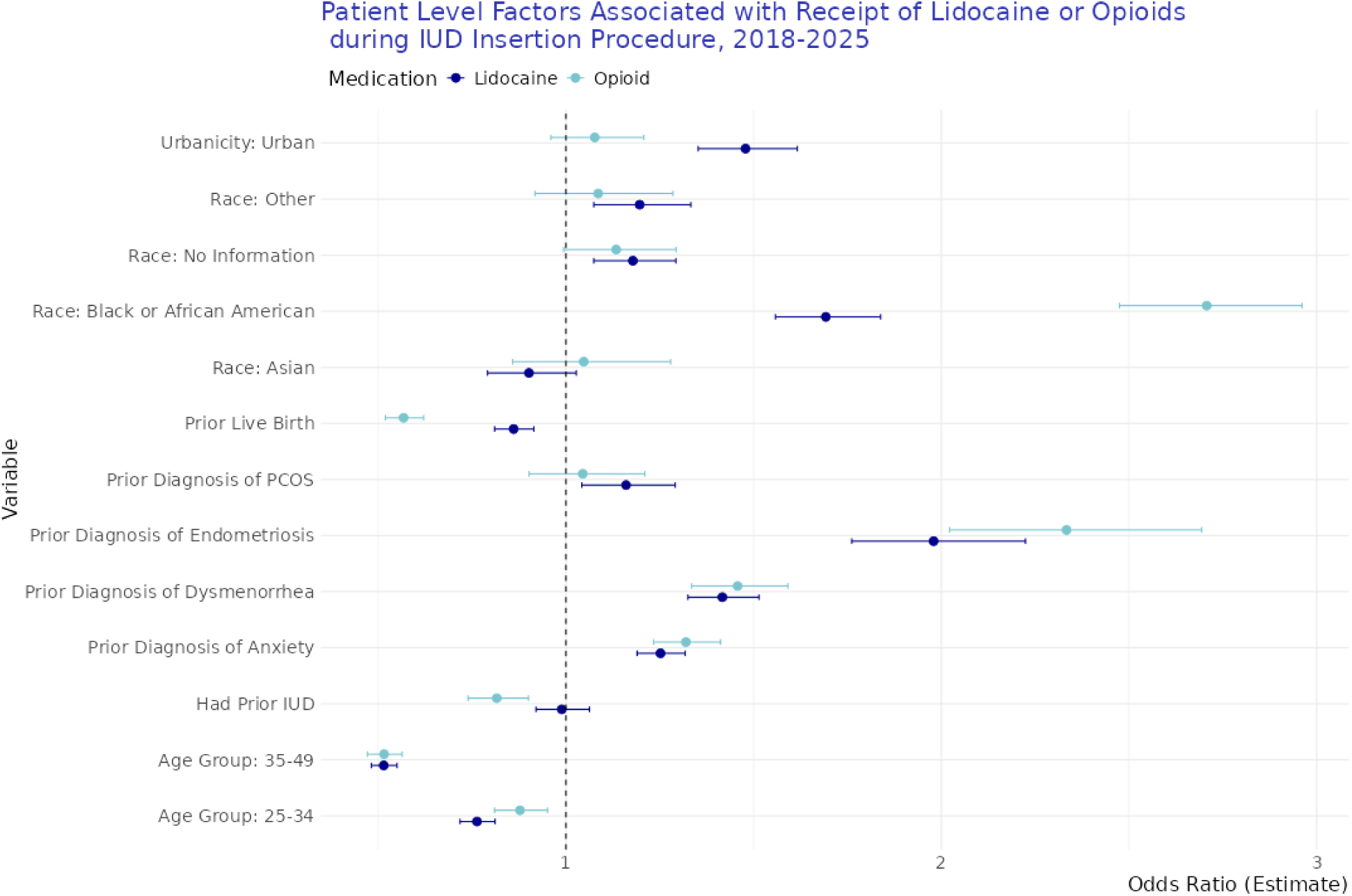
Select patient level factors from logistic regression interrupted time series models showing odds ratios of receipt of lidocaine or opioids during IUD insertion procedures. The reference level for urbanicity is rural, the reference level for race is White, and the reference level for age is 15-24. All clinical variables have a reference level of condition/diagnosis not present.

### Provider-Level Predictors of Receipt of Lidocaine and Opioids during IUD Insertions

IUD insertions with male practitioners had 10% lower odds of lidocaine receipt (aOR: 0.90 [95% CI: 0.84, 0.95]) (Figure 4, Table 2) compared to female practitioners. Compared to general practitioners (GPs, the reference group), IUD insertions where physician assistants (PAs) were involved (aOR: 2.06 [95% CI: 1.77, 2.40]) had higher odds of lidocaine administration or prescription, but no significant effect was seen for opioids. IUD insertions where medical trainees and OBGYNs were involved had higher odds of lidocaine (aOR 2.15 [95% CI: 1.79, 2.60], aOR: 1.90 [95% CI: 1.78, 2.04], respectively) and opioid receipt (aOR 2.20 [95% CI: 1.62, 2.99], aOR 3.48 [95% CI: 3.11, 3.90], respectively). IUD insertions where nurse practitioners (NPs)/registered nurses (RNs) were involved had slightly elevated odds of lidocaine receipt (aOR 1.23 [95% CI: 1.12, 1.36]), but much lower odds of opioid receipt (aOR 0.44 [95% CI: 0.35, 0.56]). IUD insertions where midwives were involved 66% lower odds of lidocaine (aOR 0.34 [95% CI: 0.25, 0.45]) and 48% lower odds of opioid receipt (aOR 0.52 [95% CI: 0.34, 0.79]).

**Figure 4.**
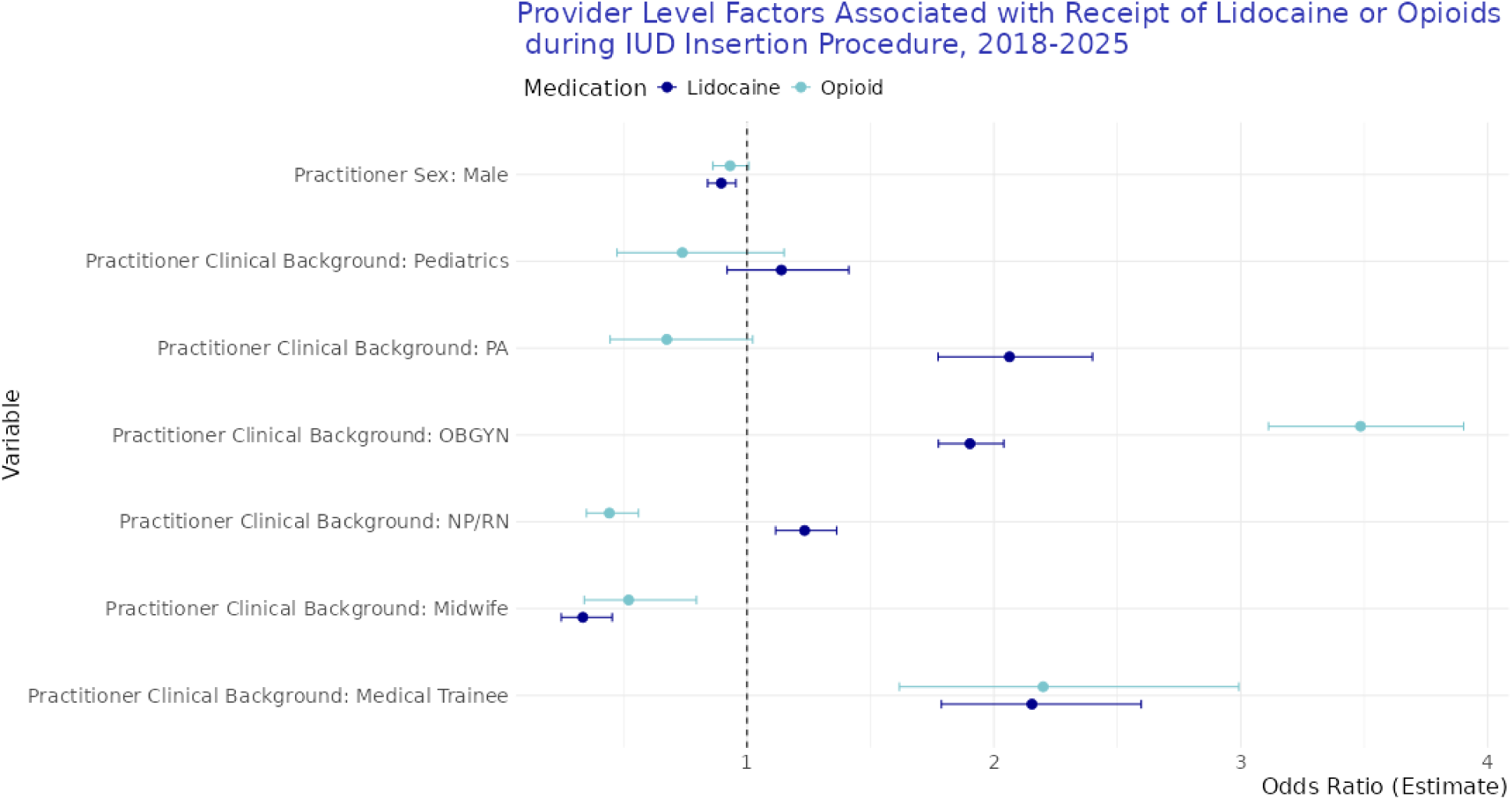
Select provider-level factors from logistic regression interrupted time series models for lidocaine and opioid administration during IUD insertion. The reference level for practitioner clinical background is general practitioner.

## Comment

### Principal Findings

There has been growing pressure to recognize the pain women experience during medical procedures, and to acknowledge how medical institutions often ignore or dismiss women’s pain.^22^ Amid growing outrage about disparities in pain management for women, IUD insertion has become a clear example of a painful procedure during which pain medication is not typically provided. Our study shows that while lidocaine administration or prescription significantly increased following the 2024 CDC guidelines, the vast majority of women – 95% – still received no pain medication for IUD insertions in 2025. Additionally, while there was a significant and immediate increase after the policy, there was no significant further monthly increase in the rate of pain medication receipt after the guidelines, highlighting that these guidelines may not be sufficient to enact a trending change, and additional discussion and momentum from professional organizations and patient advocacy may be needed.

Conditions like endometriosis, PCOS, and dysmenorrhea can involve chronic inflammation, scarring, or structural changes within the pelvis that may make IUD insertions more painful.^23–25^ We found that patients with these conditions were more likely to receive lidocaine during their IUD insertion, highlighting that clinicians may recognize the greater procedural pain experienced by these patients. Additionally, these patients may be more likely to advocate for pain relief based on their medical history, often including years of visits and dismissals from clinicians prior to their diagnoses, with the average diagnostic delay nearly 7 years for endometriosis and over 2 years for PCOS.^26,27^

We found that younger patients – specifically those aged 15-24 years – had the highest overall rate of pain medication administration or prescription during IUD insertion, reaching 6.5% in 2025. The increase in pain medication among this age group, especially beginning in 2022 (3.4% receiving pain medications), may reflect increased patient awareness and advocacy for pain medication stemming from a social media outcry about how painful IUD insertions can be.^16,17^ Younger patients may have been more attuned to this social media campaign than older patients, leading to increased self-advocacy.

### Results in the Context of What is Known

Our analysis highlighted significant differences in administration or prescription of pain medication based on the sex and clinical background of the practitioner involved in the procedure, with IUD insertions where male providers were involved having significantly lower odds of pain medication receipt. This supports research that women’s pain is often perceived to be less than that of men,^28,29^ and that provider sex may also play a role in perception of pain.^30^ Our analysis also found differences in pain medication by provider type; IUD insertions where PAs, OBGYNs, and medical trainees were involved had about twice the odds as those where general practitioners were involved of lidocaine administration or prescription, while those where midwives were involved were about a third as likely to receive lidocaine. IUD insertions where OBGYNs were involved had the highest odds of opioid receipt. These differences deserve further exploration, and may point to underlying differences in provider practices, facility characteristics, or insurance coverage.

This study reinforces and builds upon many findings from the recent study from the Veteran’s Affairs (VA) Health Care System, with about ten times the number of IUD insertions and two additional years of follow up, including formal evaluation of the impact of the CDC’s 2024 guidelines.^11^ However, the VA study included NSAIDs, and found lower rates of opioid (0.6%) and lidocaine (0.2%) administration during IUD insertions. We found higher proportions of patients prescribed or administered opioids (2.0% in 2025) and lidocaine (4.2% in 2025), which could reflect the use of both medication request and administration data in our study, as well as genuine differences between the populations in the VA or non-VA setting. Interestingly, our results showed that Black patients were significantly more likely to receive both lidocaine and opioids during IUD insertions, similar to Ware et al.’s findings.^11^ It is important to further explore racial disparities and differences in receipt of pain medication during these procedures.

### Clinical Implications

IUDs have had a complex history in North America, popularized in the 1960s and used by up to 11% of women who were using contraception.^31^ However, IUD use plummeted in the US from 7.1% in 1982 to 0.8% in 1995, likely resulting from reported risks such as pelvic inflammatory disease associated with the Dalkon Shield IUD.^32^ Since then, new generation IUDs have been introduced in the US; the copper IUD in 1988,^33^ four Levonestrogel-releasing IUDs: Mirena in 2001, Skyla in 2013, Liletta in 2015, and Kyleena in 2016,^2^ and a new copper IUD, Miudella, was granted FDA approval in February 2025.^34^ Since 2000, the popularity of IUDs has grown in the US,^35^ with 14% of women who use contraceptives aged 15-44 using an IUD from 2015-2017.^2^ IUD use varies by age, with use highest among women in their mid-late twenties.^2,35^

However, the increasing popularity of IUDs now intersects with a painful reality in medicine: a strong bias minimizing women’s pain. This bias is widespread – studies have found that women with chest pain wait 29% longer than men to be evaluated for heart attacks,^36^ and women with acute abdominal pain in the ER were up to 25% less likely than male counterparts to receive opioids for pain.^37^ However perhaps the most clear example of women’s pain being dismissed occurs in reproductive care settings. An extreme example from 2021 highlights just how much women’s pain can be dismissed: a clinic continued to perform egg retrievals despite patients expressing extreme pain, only to discover that a nurse had been stealing fentanyl and replacing it with saline, leaving patients fully unmedicated.^38^ Increasing uptake of pain medication during IUD insertions is an important way to recognize women’s pain, and to increase the acceptability of IUD insertions among those who wish to receive them.

### Research Implications

These data support a growing consensus among medical organizations that pain management during IUD insertion is insufficient. On May 15, 2025, The American College of Obstetrics and Gynecology (ACOG) updated its guidelines on pain management for in-office uterine procedures, including IUD insertion. The update recommends that clinicians offer local anesthetics for IUD insertions, including lidocaine spray, lidocaine-prilocaine cream, and paracervical block.^39^ The guidelines also highlight a trauma-informed lens to pain management, acknowledging the roles of systemic racism and patient anxiety in the experience of pain.^39^ Continued follow up is essential to evaluate how these policy guidelines are impacting patient care and provider administration of pain medication during IUD insertions.

### Strengths and Limitations

This study has several limitations, first the IUD insertion procedures included in this study were performed within a Truveta member healthcare system, and thus insertions at Planned Parenthood, community health facilities, or standalone family planning clinics would not be included. This may limit generalizability of these findings beyond IUD insertions occurring within a medical facility and underrepresent low-income and uninsured persons.^40^ Additionally, the study only captures prior births, IUDs, or comorbid conditions that occurred or were captured within a Truveta constituent health care system, leaving some medical histories incomplete for some women.

This study has several strengths, including the use of a large, geographically representative dataset which includes IUD insertions through June 2025, providing nearly real-time assessment of changes in pain medication administration during IUD insertion procedures. This study is also ten times the size of the most recent comprehensive assessment of pain medication usage during IUD insertions conducted by the VA, which included just over 28,000 insertions from 2018-2023.^11^ This study leveraged both medication request and administration data in addition to procedure data to capture pain medications prescribed, ordered or administered in the health care setting, as well as procedures for IUD insertions. Finally, this study included practitioner information, including sex and clinician background enabling practitioner-specific information to be joined to patient outcomes.

## Conclusions

While rates of pain medication administration and prescriptions during IUD procedures have increased 195% since 2018, in 2025, 95% of women receiving an IUD are still not receiving pain medication as part of the procedure. After the CDC’s updated guidelines on pain management for contraception in August 2024, pain medication usage significantly increased, however the lack of a significant trend effect since August 2024 indicates that provider behavior is not accelerating the rate of pain medication usage during these procedures. It will be essential to continue to evaluate pain medication administration during IUD insertion as more time elapses since the CDC guidelines, and to capture the impact of the 2025 ACOG guidelines on pain management for cervical procedures.

## Data Availability

The data used in this study are available to all Truveta subscribers and may be accessed at studio.truveta.com.

## CRediT statement

Conceptualization: NBM, KMG, Data curation: NBM, KMG, Formal Analysis: NBM, KMG, Investigation: NBM, KMG, NLS, PJR, BGC, DD, Methodology: NBM, KMG, PJR, Project Administration: NBM, KMG, NLS, Software: NBM, KMG, PJR, BGC, DD, Supervision: NLS, Validation: PJR, DD, Visualization: NBM, Writing – Original Draft: NBM, KMG, Writing – Review and Editing: NBM, KMG, NLS, PJR, BGC, DD.

