## Supplementary Appendix for "Evaluating Changes in Pain Medication Administration During IUD Insertion, 2018-2025"

**Table S1**: Codeset for Lidocaine and Opioid Medications*

| | Included Codes | | Excluded Codes |
|---|---|---|---|
| | **HCPCS** | **RxNorm** | **RxNorm** |
| **Lidocaine** | J2001, J2002, J2003, J2004 | 6387 | 612, 6581, 1868194, 466588 |
| **Opioids** | J3010, J0745, J0592, J0571, J0576, J1110, J2175, J2270, J1170, J0216, J1960, J2180, J1230, J2410 | 10689, 1545976, 1819, 23088, 3423, 4337, 480, 5489, 56795, 6378, 6468, 6754, 6813, 7052, 73032, 7804, 7814, 787390, 8785 | |

*Code logic uses self and descendant codes

Table S2. Codeset for Excluded Gynecological Procedures*

| Procedure Type | CPT | SNOMED | ICD10 | HCPCS |
|---|---|---|---|---|
| Biopsy | 88300, 88302, 88304, 88305, 88307, 38505, 58100, 58110, 57500, 57505, 56605, 57100, 56740, 56606, 57510, 57513, 57520 | 309283000, 50796003, 234270003, 178796002, 309284006, 386802000, 236881007, 180732009, 468629008, 287478001, 142361002 | | |
| Colposcopy | 57452, 57454, 57455, 57421, 1014191, 56820, 56821, 57420, 1014197, 1014196, 57456 | 392003006, 142513005 | | |
| LEEP | 57460, 57522, 57461 | | | |
| Hysteroscopy | 58563, 58561, 58562, 58579, 58560, 58558, 58565, 56351, 1008879, 58555, 58559 | 176878009, 142512000, 319009, 233545006 | | |
| Endometrial Ablation | 58353, 58356 | 265060005, 176874006, 149900009 | | |
| Oophorectomy | 58661, 58940, 58943, 59151, 59150, 59121, 59120 | 83152002, 149800005, 302377002, 302376006, 29672006 | | |
| Uterine Aspiration | 59812, 59820, 59821, 59830, 59840, 58120, 59841, 59857, 59856, 59855, 59852, 59851, 59850 | | 10A00ZZ, 10A03ZZ, 10A04ZZ, 10A07, 10A08 | S0199 |

*Code logic uses self and descendant codes

**Table S3:** Percent of IUD Insertions with Pain Medications Administered or Prescribed by Year, Stratified by Lidocaine or Opioid, 2018-2025

| | Percent of IUD Insertions with Pain Medications Administered or Prescribed | | | | | | | |
|---|---|---|---|---|---|---|---|---|
| **Medication** | **2018** | **2019** | **2020** | **2021** | **2022** | **2023** | **2024** | **2025** |
| Overall | 1.68 | 2.04 | 2.24 | 2.43 | 2.49 | 3.05 | 3.74 | 4.95 |
| Lidocaine | 1.20 | 1.61 | 1.83 | 2.01 | 2.17 | 2.57 | 3.21 | 4.22 |
| Opioid | 0.84 | 1.12 | 1.14 | 1.09 | 1.11 | 1.49 | 1.74 | 2.04 |

**Table S4:** Percent of IUD Insertions with Pain Medications Administered or Prescribed by Year, Stratified by Age, 2018-2015

| | Percent of IUD Insertions with Pain Medications Administered or Prescribed (Total) | | | | | | | |
|---|---|---|---|---|---|---|---|---|
| **Age Group** | **2018** | **2019** | **2020** | **2021** | **2022** | **2023** | **2024** | **2025** |
| 15-24 Years | 2.08 | 2.47 | 2.42 | 2.81 | 3.41 | 3.97 | 5.35 | 6.54 |
| 25-34 Years | 1.63 | 2.14 | 2.34 | 2.64 | 2.45 | 3.29 | 3.77 | 5.28 |
| 35-49 Years | 1.43 | 1.63 | 1.87 | 1.93 | 1.96 | 2.20 | 2.84 | 3.74 |
